# Impact of a multi-disease integrated screening and diagnostic model for COVID-19, TB, and HIV in Lesotho

**DOI:** 10.1101/2022.12.20.22283748

**Authors:** Bulemba Katende, Moniek Bresser, Mashaete Kamele, Lebohang Chere, Mosa Tlahali, Rahel Milena Erhardt, Josephine Muhairwe, Irene Ayakaka, Tracy R Glass, Morten Ruhwald, Bram van Ginneken, Keelin Murphy, Margaretha de Vos, Alain Amstutz, Mathabo Mareka, Sekhele Matabo Mooko, Niklaus D. Labhardt, Klaus Reither, Lucia González Fernández

**Affiliations:** SolidarMed, Partnerships for Health, Maseru, Lesotho; Swiss Tropical and Public Health Institute, Allschwil, Switzerland; University of Basel, Basel, Switzerland; Butha-Buthe District Health Management Team, Butha-Buthe, Lesotho; Mokhotlong District Health Management Team, Mokhotlong, Lesotho; FIND, the global alliance for diagnostics, Geneva, Switzerland; Diagnostic Image Analysis Group, Radboud UMC, Nijmegen, The Netherlands; Department of Infectious Diseases and Hospital Epidemiology, University Hospital Basel, Basel, Switzerland; CLEAR Methods Center, Division of Clinical Epidemiology, Department of Clinical Research, University Hospital Basel, University of Basel, Basel, Switzerland; National Reference Laboratory, Ministry of Health of Lesotho, Maseru; SolidarMed, Partnerships for Health, Lucerne, Switzerland

**Keywords:** SARS-CoV-2, COVID-19, HIV, Tuberculosis, integrated TB/COVID-19, sub-Saharan Africa, Lesotho

## Abstract

**Introduction:** The surge of the COVID-19 pandemic challenged health services globally, and in Lesotho, the HIV and tuberculosis (TB) services were similarly affected. Integrated, multi-disease diagnostic services were proposed solutions to mitigate these disruptions. We describe and evaluate the effect of an integrated, hospital-based COVID-19, TB and HIV screening and diagnostic model in two rural districts in Lesotho, during the period between December 2020 and August 2022.

**Methods:** Adults and children above 5 years attending two hospitals were screened for COVID-19 and TB symptoms. After a positive screening, participants were offered to enroll in a service model that included clinical evaluation, chest radiography, SARS-CoV-2, Xpert MTB/RIF Ultra and HIV testing. Participants diagnosed with COVID-19, TB, or HIV were contacted after 28 days evaluate their health status, and linkage to HIV or TB services.

**Results:** Of the 179160 participants screened, 6623(37%) screened positive, and 4371(66%) were enrolled in this service model, yielding a total of 458 diagnoses. One positive rapid antigen test for SARS-CoV-2 was found per 11 participants screened, one Xpert-positive TB case was diagnosed per 85 people screened, and 1 new HIV diagnosis was done per 182 people screened. Of the 321(82.9%) participants contacted after 28 days of diagnosis, 304(94.7%) reported to be healthy. Of the individuals that were newly diagnosed with HIV or TB, 18/24(75.0%) and 46/51(90.1%) started treatment. This service showed no difference in the detection of new HIV and TB cases when compared to other hospitals, where no such integrated service model was provided.

**Conclusion:** This screening and diagnostic model successfully maintained same-day, integrated COVID-19, TB, and HIV testing services through different COVID-19 incidence periods in a resource-limited context. There were positive effects in avoiding diagnostic delays and ensuring linkage to services, however, efficiencies were contingent on the successful adaptation to the changing environment.

## Introduction

The spread of the severe acute respiratory syndrome coronavirus 2 (SARS-CoV-2) and the coronavirus disease 2019 (COVID-19) pandemic negatively impacted health services delivery in most countries, even where health systems were thought to be well-established(1–4). At the same time that the pandemic created a significant increase in morbidity and mortality globally, health facilities across the globe shut down or restricted their activity to the provision of essential services. Factors such as infection control risks, lack of health workers or personal protective equipment(5) led to a substantial reduction in access to primary health care worldwide(6). In countries with high TB and HIV prevalence, the COVID-19 pandemic reversed years of progress in the control of these 2 conditions(7,8).

Although the African continent reported relatively low COVID-19 cases, compared to other regions in the world, the impact of the pandemic on health service delivery was still very significant. Access to health facility–based services was interrupted as a result of 1) health workers shortages due to illness, self-quarantine, or strikes, 2) health facilities repurposed for COVID-19 treatment, covering minimum services, or closed completely, and 3) clients’ reluctance to attend to visits, due to fear of increased risk of COVID-19 exposure, or decreased availability of public transportation(6,9,10). A review by Gizachew A. Tessema *et al*. reported substantial reduction in access to essential and general health services across the African continent(11), and in South Africa, a study in the KwaZulu Natal province reported an estimated 47.6% and 46.2% decrease in HIV testing and anti-retroviral therapy initiation respectively(12).

Lesotho is a country in the sub-Saharan region with over 2 million population(13). The estimated TB incidence sits at 614%, the HIV prevalence is 20.9%, and there is approximately 55% TB/HIV coinfection(14–16). Similar to other settings, the health system and health services in Lesotho were similarly affected by the COVID-19 pandemic(17–21). The first COVID-19 case was reported in May 2020, and up to December 2022, approximately 34,490 COVID-19 cases and 706 COVID-19 related deaths have been reported. Although cases have been recorded continuously since May 2020, three significant waves were described, with high incident rates in December 2020, June to September 2021, and December 2021(22). To reinforce the efforts against the spread of SARS-CoV-2, the government of Lesotho set up a public health strategy that regulated social norms to ensure physical distance between people. A COVID-19 Risk Determination and Mitigation Framework was created as a 5-colour scale, regulating daily life activities, depending on the COVID-19 reproductive number (R0) determined regularly. The scale of colors varied from green, blue, purple, yellow, and red. Green color was associated with R0 <1 and red color was associated with R0 > 2.5. Typically, at each subsequent level, social measures became more stringent, and included mandates, such as staying home, avoiding crowds, or closure of business and borders (23–28).

In this context, the project named *Mitigation strategies for communities with COVID-19 in Lesotho* -MistraL-(29) was conceived as an initiative to expand COVID-19 diagnosis and integrate TB and HIV testing in the districts of Butha-Buthe and Mokhotlong, situated in the northeast part of the country (figure 1). From December 2020 to August 2022, MistraL reinforced the health system response (table 1), contributing to aspects such as: 1) support to the COVID-19 response coordination at district and national level; 2) addition of resources to improce health facilities infrastructure and provision of medical equipment; 3) integration of COVID-19/TB/HIV testing and linkage model; 4) addition and training of health workers for provision of services; and 5) implementation of research activities, including SARS-CoV-2 RTT tests validation in nasopharyngeal and nasal samples, or use and validation a of computer-aided diagnostic software for COVID-19 (CAD4COVID).

In recent years, there has been an increasing interest to understand whether and how integrated screening and testing services could provide access to prompt COVID-19 diagnosis and management, while maintaining diagnostic and linkage services for TB and HIV, in an epidemic context(30–32). Nonetheless, to date, there have been few studies demonstrating feasibility or wider impact of such services in sub-Saharan Africa(33–35). The objective of this study is to describe and assess the effect of a pragmatic COVID-19/TB and HIV integrated screening and testing service model for adults and children older than 5 years in two remote referral hospitals in Lesotho.

## Methods

### Context

We implemented this screening and diagnostic model at the St Charles Missionary Hospital Seboche and the Mokhotlong Government District Hospital, in the districts of Butha-Buthe and Mokhotlong. These districts are characterized by mostly rural settings with an estimated combined population of 250,000 people, who are mainly subsistence farmers, mine workers or construction and domestic laborers who work in neighboring South Africa. Each district has only one single mid-size town: Butha-Buthe with ca. 25,000 inhabitants and Mokhotlong with ca. 10,000 inhabitants. The remaining population lives in villages scattered over a mountainous area of 5,842 km^2^. Health services in these two districts are provided through a network of clinics and hospitals. In Butha-Buthe district health care is available thought 10 nurse-led rural health centers, 1 missionary hospital, and 1 governmental hospital. In Mokhotlong services are available thought 9 nurse-led rural health centers, and 1 governmental hospital.

### Integrated TB/COVID-19 and HIV screening and testing model

A nurse-led, multi-disease, integrated screening and diagnostic model was established, with the aim to provide same-day, one-stop shop diagnosis and clinical evaluation. After the diagnostic procedures were done, the nurses would manage cases themselves, or refer to other hospital staff, according to stablished clinical algorithms (figure 2). This service was continuously adapted to the needs and diagnostic standards, driven by a fast-changing context.

### Screening, eligibility, and enrolment

Adults and children seeking health services, visitors, and staff attending daily work, were pre-screened by trained TB/COVID-19 lay screeners. Pre-screening took place in specially built structures by the gates of the two hospitals and focused on the investigation of history of close contact with a COVID-19 case or the presence of at least one sign or symptom of TB or COVID-19. The initial evaluation included: fever measured (≥38 °C, forehead or reported), cough of any duration, pronounced tiredness, shortness of breath, sore throat, muscle or body pain, diarrhoea, loss of taste/smell, weight loss, night sweat. We defined close contact with a (confirmed or probable) COVID-19 case, as a contact within one meter for more than 15 minutes, or direct physical contact with a (probable or confirmed) case, or direct care provided to a patient with (probable or confirmed COVID-19) disease without using proper personal protective equipment. A significant contact of TB was determined using the standard definition(36). We defined positive pre-screening as the presence of at least one sign or symptom, or a history of close contact with a COVID-19 or TB case. People who pre-screened positive and were 5 years or older, were referred to a nurse, who determined eligibility to enroll in this diagnostic model, explained the testing procedures, and obtained informed consent.

Once the participant was enrolled, the nurse took medical history focusing on COVID-19, TB, and HIV, measured BMI, body temperature, and performed a clinical examination to assess the severity of respiratory symptoms. We defined mild, moderate and severe clinical presentation as follows: 1) mild disease included the presence of COVID-19 signs and symptoms, without hypoxia or dyspnea; 2) moderate disease included the presence of signs and symptoms of pneumonia (fever, cough, dyspnea, tachypnoea) or an oxygen saturation ≤ 94%, with no signs of severe pneumonia; and 3) severe disease included the presence of at least one of the following: altered mental status, oxygen saturation < 94% or signs of central cyanosis, shortness of breath or difficulty breathing, fast breathing (5-9 y ≥ 30 breaths/min; ≥10 y ≥ 20 breaths/min; and ≥18 y ≥ 22 breaths/min), systolic blood pressure <100 mmHg in adults and general danger signs in children. For severely ill participants, the nurse referred them to a physician in the hospital immediately after the clinical examination and patients followed the integrated diagnostic procedures only if the physician judged that these would benefit them. Patients with mild or moderate symptoms at presentation remained in the study and were offered all the clinical procedures. Enrolled participants had a digital posterior-anterior chest Xray done and analyzed using artificial intelligence-powered, computer-aided detection software for TB (CAD4TB version 6; Delft Imaging Systems, the Netherlands). Those with a result score of 50 or above followed investigations for TB(37).

### TB, COVID-19, and HIV diagnosis

Diagnosis of SARS-CoV-2 infection was done using RDTs or PCR, in different combinations that varied along the implementation period, adapting to the enrolment of different studies, and the evolving diagnostic recommendations for SARS-CoV-2 in Lesotho. We used the STANDARD Q COVID-19 Antigen test (SD Biosensor, Republic of Korea) with a nasal or a nasopharyngeal swab, and PCR (ABI 7500 Real-Time PCR platform, Applied Biosystem, USA and Xpert Xpress SARS-CoV-2 Cepheid, USA). All participants included in this study had at least one nasopharyngeal SARS-CoV-2 RDT result. Participants who presented with presumptive TB symptoms or a CAD4TB score > 50 were requested to submit sputum for TB diagnostic testing using the Xpert MTB/RIF Ultra (PCR GeneXpert, Cepheid, Sunnyvale, CA, USA). Participants who were requested to submit sputum watched a video describing how to produce a quality sputum before they attempted to produce the sample. For HIV diagnosis, we followed standard of care in Lesotho, that included a first assessment using an HIV risk screening tool to determine HIV testing eligibility. If participants were found to be high risk using the HIV testing screening tool, they were tested following the Lesotho HIV guidelines that used standard blood based RDTs(38). For the purpose of this study, we defined a COVID-19 case as an individual with a positive result in either RDT (nasopharyngeal or nasal swab) or PCR (nasopharyngeal swab). TB diagnosis was defined as a positive Xpert MTB/RIF Ultra result in sputum, and an HIV diagnosis was defined according to Lesotho standard testing algorithm.

### Follow up after 28 days after testing

To measure the effect of this integrated multi-disease diagnostic service, in terms of linkage to COVID-19 curative services, TB and HIV treatment initiation, all enrolled participants that had at least one TB, COVID-19 (RDT or PCR) or HIV positive result, were followed up in a window of time around 28 days after enrolment. A study nurse attempted to contact each participant or their next in kin, through a phone call. In the case of participants diagnosed with COVID-19, we collected information on whether they were “healthy” (had recovered from symptoms, after the episode), “hospitalized” (still admitted due to the condition) or deceased. For participants that were newly diagnosed with TB or HIV, we asked whether they had started treatment after diagnosis and checked hospital records (TB and HIV registers) at the two hospitals.

### Ethical Consideration

All procedures were carried out in line with the ethical standards laid out in the Declaration of Helsinki(39). Participants that enroll in this diagnostic model received information on the clinical procedures in Sesotho and gave written informed consent and those who were illiterate gave consent by thumbprint and a witness signature. Parents or guardians of all participants below 18 years signed an informed consent form, while participants between 7 and 17 years additionally signed an assent form. This project was approved by the Lesotho Ministry of Health Ethics Committee (ID 107-2020) and by the Ethic Committee Switzerland (EthikkommissioNordwest-und Zentralschweiz(EKNZ) AO_2020-00018).

### Data collection and analysis

Clinical data was collected by trained nurses using the Open Data Kit (ODK) software(40). Information that could not be obtained on the same day, such as Xpert MTB/RIF Ultra and SARS-CoV-2 PCR results, was uploaded to ODK as soon as was available. Routine data quality checks were done regularly. To compare TB and HIV indicators across hospitals we obtained data from the Lesotho District Health Information System 2(DHIS 2)(41). Information about social restrictions, COVID-19 policies modification, and COVID-19 national response in Lesotho was compiled using the Lesotho Ministry of Health sources. Descriptive statistics were used to characterize the study sample. Median and interquartile range (IQR) were calculated to describe continuous variables, and frequencies and percentages for categorical variables. Statistical analysis was performed using Stata (version 16.1, College Station, Tex: StataCorp LP, 2007).

## Results

Of the 179160 adults and children that were pre-screened, a total of 6623 (37%) had at least one COVID-19 symptom or history of close contact with a COVID-19 or TB case. Amongst those who pre-screened positive, a total of 4371 (66%) were enrolled in the integrated TB/COVID-19/HIV testing procedures. The remaining 2252 (34%) participants that did not enrolled despite presenting symptoms were: 1) children below 5 years, 2) unaccompanied children between the age of 7 and 17 years, 3) patients presenting in the weekends, when the service was not available, 4) patients with severe symptoms that required immediate medical attention, or 5) people who refuse to enroll in this service. Table 2 provides an overview of the baseline characteristics of enrolled participants. A total of 2468 (61,7%) were adult women and 369 (8.4%) were children and adolescents between the age of 5 and 17 years. Most of them presented with at least one symptom, most commonly cough, or pronounced tiredness. The median time from symptoms onset was 3 (IQR 2 -7) days, more than 98% presented with mild or moderate symptoms and had an oxygen saturation ≥ 95%. A total of 1768 (44.1%) adults had overweight or obesity (BMI ≥25 Kg/m2), 1058 (27.8%) had one elevated blood pressure measurement (≥140/90 mmHg), and 321 (8.0%) reported current tobacco use. A total of 125 (3.7%) adults and 2 (0.6%) children reported a recent TB contact, while 111 (3.3%) adults and 6 (1.9%) children reported a recent COVID-19 contact. A total of 814 adults (20.3%) and 28 children (7.6%) reported to live with HIV.

### Integrated COVID-19/TB and HIV screening and testing model

A summary of the diagnostic results is available in table 3. In terms of SARS-CoV-2 testing, a total of 4355, 2421 and, 2664 nasopharyngeal RDT, nasal RDT, and nasopharyngeal PCR tests were done. With regards to TB investigations, a total of 4371 chest x-rays were performed, and overall, 884 (29.3%) participants had a CAD4TB score > 50. For the 2419 participants who were eligible for sputum collection, 1454 samples were collected and tested with Xpert MTB/RIF Ultra. Most common reasons for not collecting sputum included patient refusing (377; 15,8%) or unable to produce a sample (263; 10.8%). A total of 104 people was tested for HIV. Overall, a total of 458 new diagnoses were made, of which 383, 51 and 24 were COVID-19, TB, and HIV cases, respectively (table 3). Newly found cases were mainly diagnosed among adults. Only 16 (4.3%) of the COVID-19 cases and 1 (1%) of the TB cases were diagnosed in children and adolescent participants, and this model did not find any new HIV diagnosis amongst them.

With regards to the clinical outcomes after four weeks of diagnosis, of the 458 people who were diagnosed with either COVID-19, TB or HIV, a total of 321 (82.9%) participants, or their relatives could be reached. Of those, 304 (94.7%) reported that they were healthy, whereas 5 (1.3%) reported to be ill or hospitalized (four people diagnosed with COVID-19 and one person who was HIV positive). We recorded 12 (3.7%) participants who died in this period, however we did not record the possible causes of death. Of the individuals that were newly diagnosed with HIV or TB, a total of 18/24 (75.0%) and 46/51 (90.1%) had started ART and TB treatment respectively, during this period (table 3).

The yield of this integrated testing model in terms of aditional HIV, TB, and COVID-19 diagnosis across periods of different COVID-19 incidence is displayed in figure 3. Periods 1 and 4 (end of 2020 to mid-February 2021, and end of November 2021 to end of February 2022), are characterized by high SARS-COV-2 RDT positivity rates (20-40%), during very short periods of time (distinct COVID-19 waves). Periods 3 and 5 (mid-June to end of November 2021, and end of February to end of August 2022), are marked by lower SARS-COV-2 RDT positivity rates across longer periods of time, indicating ongoing, low level of community transmission. During period 2 (mid-February 2021 to Mid-June 2021), there were almost no COVID-19 cases diagnosed at our sites. During periods of high COVID-19 incident cases (1 and 4), when study staff had to cope with a larger number of enrolled participants, the number of TB and HIV additional cases found was irregular. Whereas the periods of lower COVID-19 incident cases allowed for a higher number of new TB and HIV diagnosis.

### Comparison performance of TB and HIV services with similar hospitals

Figure 4 shows trends of HIV and TB new notifications, as well as the ART initiations, reported in six hospitals Lesotho during the period of October 2020 to March 2022, using the Lesotho DHIS-2 reporting system. Portrayed sites include the two study sites (Seboche Mission Hospital and Mokhotlong Hospital), compared to Paray, St. James, Ntseke, and Tellebong hospitals, where COVID-19 testing was conducted but not integrated with TB and HIV services. All hospitals maintained HIV and TB services similarly during this period and the Seboche and Mokhotlong hospitals, where this diagnostic model was implemented, seemed to perform quite similarly.

## Discussion

This observational study aimed to describe and assess the impact of an integrated COVID-19, TB and HIV screening and testing service model in two hospitals in rural Lesotho, during the period between December 2020 and August 2022. Our setting was characterized by an overall low incidence of COVID-19 cases, a volatile context, and social restrictions in continuous revision. Nonetheless, this service operated during two distinct COVID-19 waves and two other periods when COVID-19 incident cases were continuously low. This pragmatic diagnostic service model provided integrated diagnostic services to 4,371 adults and children, who most frequently presented with non-severe symptoms, and previous contact with a TB or COVID-19 case was unfrequently reported. The overall diagnostic yield, including COVID-19, TB, and HIV, was 458 new cases, most frequently found in adults; only 17 (3.7%) of them happened in children and adolescents.

It is almost certain that in this study we underestimate the number of COVID-19 and TB cases diagnosed. In the case of COVID-19 cases, a PCR test was used as the gold standard for diagnosis during most part of 2021. Later on, diagnosis was based on the result of the SARS-CoV-2 STANDARD Q COVID-19 Ag test, that has shown relatively low diagnostic performance in our setting (42). Anecdotically, a number of the 2252 (34%) eligible adults and children that presented with symptoms but did not enroll in this service model despite being eligible, were still tested for COVID-19 by the study staff, especially in periods were hospital health staff in other departments was scarce, however, these individuals are not included in this analysis. With regards to the definition of TB case in this study, we report only positive Xpert MTB/RIF Ultra results in the sputum of participants, from whom we obtained a sample. In our case, sputum samples could not be obtained for one third of the participants, mainly due to refusals or inability to produce them. Additionally, for this analysis, we did not consider a clinical TB diagnosis made by clinicians once a symptomatic patient was referred to further evaluation by hospital staff.

To interpret these findings, it is also important to consider broader contextual factors that played a role. One of them is related to the variation of circulating SARS-CoV-2 virus (Omicron variant) and the lower diagnostic sensitivity observed using RDTs(43,44). In our setting, this could have led to a lower detection or cases in symptomatic people seeking services. A second factor could be related to the fluctuating access to hospital services. It is probable that the changes imposed by the social restrictions, the availability of service providers (themselves ill, in quarantine or isolated due to a recent contact), or the fear to acquire a COVID-19 infection in the hospital, were important deterrents for symptomatic people to use hospital services.

Of the 321 (82.9%) participants that were successfully reached after four weeks of the diagnosis, 304 (94.7%), 18/24 (75%), and 46/53 (86.8%) were in good health, had started ART and TB treatment respectively during this period. We recorded 12 (3.6%) participants deaths, however, around one in six participants with a diagnosis could not be contacted in the follow up period, therefore these results need to be interpreted with caution. Using the total number of 4371 enrolled participants as a benchmark, we calculate that we found one SARS-CoV-2 rapid antigen positive per 11 people screened, one Xpert MTB/RIF Ultra positive test per 85 people screened, and 1 HIV new diagnosis was found per 182 people screened.

Surprisingly, we found no clear evidence to suggest that this resource-intense screening and testing model had a significantly different impact in sustaining access to TB and HIV diagnostic and treatment services, compared to similar hospitals in Lesotho, where this diagnostic model was not in place, when referring to the Lesotho TB and HIV DHIS-2 reports. During the analyzed period of October 2020 to March 2022, the number of HIV new diagnosis, ART initiations, and TB case notifications in the six hospitals seem quite similar. Additionally, the results from our study suggest that this diagnostic model had little impact on pediatric and adolescent COVID-19, TB and HIV case finding in participants who were between the age of 5 and 18 years. On the other hand, the overall TB and HIV linkage to care rates within four weeks of diagnosis were higher than reported in similar experiences(45–48), however when comparing to global targets, this model fell short in ART initiation rates (75% vs 95%), and met the TB treatment initiation target (90% vs. 90%)(49,50).

As the COVID-19 pandemic surged, and lockdowns led to the cancellation of essential health services, global health actors advocated to adapt services that integrated TB, HIV and COVID-19 activities in sub-Saharan Africa(29,42–44). This established an emerging body of evidence that evaluates the experiences and successes in the integration of services. A study in Uganda, using qualitative methods(34), reported that facilitators to integrate COVID-19 and TB services included availability of focal health workers who were solely responsible for this task, and availability of standard procedures and data collection tools. In this setting, inconsistent supply of protective gear and fear of contracting COVID-19 decreased the efficiency of the service. A study in Ethiopia reported a sharp reduction of TB service indicators in Addis Ababa, partially mitigated with the use of digital health technology to screen for TB, however, their results were not conclusive, due to lack of patient-level data(33). Another similar experience in Niger and Guinea(54), found that out of the 863 individuals enrolled, 61 (7%) tested positive for COVID-19 and 43 (4.9%) were diagnosed with TB. These results are comparable to our COVID-19 notifications in Lesotho (383, 8.8%), however, the investigators in Niger and Guinea found a higher number of TB cases than in our setting [35 (4.6%) in Guinea, 8 (7.6%) in Niger, and 51 (1.1%) in Lesotho]. Reports of pragmatic integrated experiences aiming to sustain HIV testing and linkage to treatment amid the COVID-19 epidemic across sub-Saharan Africa are very scarce. A systematic review evaluating the impact of the COVID-19 pandemic on accessing HIV services in South Africa, and an evaluation of performance of services in primary health clinics(55,56) revealed that there were significant decreases in HIV testing, positive HIV tests, and ART initiation at public health facilities. However, the private facilities had maintained their activity similarly to pre-pandemic levels, confirming a disparity in health access between the private and public sectors. In our service, the routine use of the HIV testing screening tool targeted individuals who were most at risk of a recent infection and yet, one in five people tested positive for HIV. However, it is important to note that in our setting performing HIV testing depended greatly on the availability of time that the study nurses had, while being burdened with other tasks. In periods of sharp COVID-19-related overload, created by an increase of influx of individuals looking to be tested, participants who were also eligible for HIV testing were referred to the routine hospital HIV testing services. This was a result of an effort to maintain the function of the COVID-19 diagnostic services and the quality of the HIV testing service. This typically happened in a different hospital department and could have decreased the efficiency to find new HIV cases. Similarly to other published studies in sub-Saharan Africa and other regions of the world(33– 35,54,57–59), our integrated screening and diagnostic model proved to be a resource intensive service, that had to adapt to contextual factors continuously. To ensure availability and continuity of the service, we made significant investments in the deployment, protection, training, and support of health staff.

Our study had various limitations. A major weakness is that we report on a hospital-based screening and testing model, that served the population that voluntarily reached and enrolled in the service. Therefore, our results refer to a selected population. By the end of 2021, SARS-CoV-2 ag-RDTs became available at community level, through local retailers and primary health clinics. As capacity for rapid testing for COVID-19 expanded, the population living in these districts could choose where and how receive this service. Secondly, complementing our study with qualitative approaches could have increased the efficiency, and acceptability of the integrated diagnostic service, and explored the level and consequences of stigma related to COVID-19 in our setting. However, these aspects were explored from the perspective of the community in the two districts and will be reported in other studies. Lastly, an economic analysis of this model could shed light on important questions related to health system-related costs and patients-related costs. Despite these limitations, this is the first study evaluating the effect of an integrated, COVID-19/TB and HIV diagnostic model in sub-Saharan Africa, covering a period of 22 months and reporting on disaggregated characteristics and outcomes in adults and children above 5 years.

### Conclusion

This study shows that integrated testing for TB/COVID-19 and HIV is feasible in resource-constraint, routine programmatic settings. Our findings provide information to support health system managers and policy makers in the design of services that integrate COVID-19 aspects. Continuous surveillance of COVID-19 incident cases, availability of health staff, and routine monitoring of diagnostic yields are important aspects to adapt these services and find efficiencies in such rapid-evolving contexts.

## Data Availability

All data will be made available as described in the PLOS Data Policy

## Declarations

### Ethics approval and consent to participate

This project was approved by the Lesotho Ministry of Health Ethics Committee (ID 107-2020) and by the Ethic Committee Switzerland (EthikkommissioNordwest-und Zentralschweiz (EKNZ) AO_2020-00018)

### Consent for publication

All authors consent to the publication of this manuscript.

### Availability of data and materials

All data generated or analyzed during this study are included in this published article [and its supplementary information files]. All articles used for the review are in the references section.

### Author Contributions

BK, KR, and LGF conceptualized the study and design. BK, MB, RME, TG performed references screening, data collection and summarization. BK, KR, and LGF drafted the manuscript. All authors reviewed the results and approved the final version of the manuscript.

